# How can we increase notification of exposure to STIs between gay and bisexual men?: intervention development using stakeholder engagement, qualitative methods and behavioural science

**DOI:** 10.1101/2021.05.27.21257903

**Authors:** Paul Flowers, Makeda Gerressu, Julie McLeod, Jean McQueen, Gabriele Vojt, Merle Symonds, Melvina Woode-Owusu, Claudia Estcourt

## Abstract

**Rationale:** The first key step in contact tracing for sexually transmitted infections (STIs) is to *notify* recent exposed sex partners. Gay and bisexual men and other men who have sex with men (GBMSM) bear a high burden of STIs and one-off partners contribute disproportionately to community transmission, posing a particular challenge to contact tracing. Here we explore and theorise the barriers and facilitators of GBMSM telling their ‘one-off’ sexual partners about their exposure to STIs.

**Design:** Using focus groups with diverse GBMSM from Leeds, Glasgow, London and on-line (n=28) we used a multi-level approach to intervention development to enhance contact tracing. This framework included initial stakeholder engagement; deductive thematic analysis to identify key barriers and facilitators to contact tracing with one-off partners; the use of the theoretical domains framework (TDF) to theorise these barriers and facilitators and subsequently the use of the behaviour change wheel (BCW), incorporating the behaviour change technique taxonomy (BCTT), to suggest intervention content to enhance the key step of notifying partners; and final stakeholder input to ensure this content was fit for purpose and satisfied the APEASE criteria.

**Results:** In relation to the TDF, the barriers and facilitators primarily related to ‘beliefs about consequences’. Having used the BCW and further stakeholder engagement, our final intervention recommendations related to focussed efforts to change the culture and corresponding norms and social practice of notifying sex partners about the risk of infection in GBMSM communities. This could usefully be achieved through dedicated community engagement and partnership work, through focussed mass and social media interventions twinned with focussed peer-led work to normalise and destigmatise contact tracing.

**Conclusion:** Through systematically working with key stakeholders, GBMSM communities and using a range of tools from the behavioural sciences, we have developed a suite of evidence-based and theoretically informed intervention content which, if developed further, could enhance GBMSM’s willingness to notify sex partners about their risk of infection.

## Background to contact tracing for sexually transmitted infections (STIs) – partner notification (PN)

COVID-19 has highlighted the centrality of contact tracing to infectious disease prevention. Although it is within the context of COVID-19 that many people have come to understand contact tracing, it has been a central tool in the control of STIs for decades. STIs continue to be a significant public health concern globally because of increasing new diagnoses, complications and long-term health problems if left untreated, and the cost to healthcare services (Health Matters, 2019; WHO, 2019).

In relation to STIs, contact tracing is typically referred to as partner notification (PN). Definitions of PN are wholistic and encompass informing sexual partners of their possible exposure and providing testing and treatment as appropriate as well as advice for future prevention (McClean et al., 2012). In this way, PN facilitates timely access to testing and health care to affected individuals but also holds a public health role in interrupting chains of transmission and reducing the spread of STIs (World Health Organisation (WHO)/UNAIDS, 1999; Ferreira et al., 2013). Although there are many actors, actions and settings involved in the whole process of PN (see Flowers et al., *in preparation*) here, we only focus on the “gateway step” in which GBMSM notify their partners of their exposure to an STI. Elsewhere we report on a more comprehensive approach to developing an intervention to improve PN (Flowers et al., 2021).

### GBMSM and changes in STIs

Recovery from the HIV pandemic has been associated with the re-emergence of gonorrhoea and syphilis among gay and bisexual men who have sex with men (henceforth, GBMSM) (WHO, 2019). In England, GBMSM accounted for 47% of gonorrhoea and 75% of syphilis diagnoses in 2018, with respective diagnoses rising by 643% and 236% since 2009 (PHE, 2019). Changes in sexual behaviour including increases in condomless sex, perhaps as a response to PrEP, likely contributed to these figures (PHE, 2019). Further, concerns about gonorrhoea have arisen globally due to rising antimicrobial resistance (Wi, Lahra et al., 2017).

### PN and MSM

Achieving good outcomes of contact tracing in GBMSM is challenging. GBMSM tend to report greater numbers of sexual partners and a higher frequency of casual and anonymous sexual encounters than most heterosexuals (PHE, 2014; Mercer et al., 2016). These factors impact on the ability and decision to engage with PN (van Aar, 2012; Wang, 2016). The increasingly popular use of geosocial networking (GSN) apps among GBMSM to meet sexual partners has also changed the fundamentals of PN when contact details might solely be shared on an app and users may not use stable or unique profile names (Contesse et al., 2019). It is worth noting however that it is likely these digitally mediated ways of meeting are more likely to facilitate PN than meeting through public sex environments for example (e.g., see Flowers et al., 1999; Flowers et al., 2000)

### PN and partner type

When diagnosed with an STI, people’s abilities to contact sexual partners and the motivation to do so relate to the kind of relationship that connects them. Working with a range of evidence sources, to assist with considering the tailoring of PN approaches to relational contexts, we developed a typology of sexual partner types (‘Established partner’, ‘New partner’, ‘Occasional partner’, ‘One-off partner’ and ‘Sex worker’) (Estcourt et al., 2021). Our evidence synthesis suggested that PN was most challenging with ‘one-off partners’. These partnership contexts are most likely to face issues with accurate contact details, most likely to be challenging because of the lack of emotional connection between partners facilitating care of the other, and further because, as no future sex is planned, there is no personal benefit in notifying partners to avoid reinfection. From the service perspective, this kind of relationship type can be particularly labour intensive and creates an additional service burden as they are the least likely to be easily contacted (Ruscher et al,. 2019).

### Behavioural Theory

To increase notification amongst GBMSM and their one-off partners, we have drawn upon key tools from the behavioural sciences that encompass understanding and theorising underlying reasons for behaviour, and then moderating important mechanisms likely to elicit behaviour change.

Michie and colleagues (2011) developed a theory and evidence-based model of behaviour change, the behaviour change wheel (BCW) approach, to systematically develop behaviour change interventions. The BCW incorporates various analytical tools which all share a background in terms of being created from the synthesis of previous expert knowledge, previous theories, and wider approaches to behaviour change. The tools have been developed through consensus building and span health psychology and an array of relevant inter-disciplinary perspectives. As such, the BCW approach and its tools can be considered as all encompassing ‘meta-frameworks’.

Together, these meta-frameworks provide a common language for describing intervention components across a range of levels from the macro to the micro (i.e., ‘policy categories’, ‘intervention functions’, ‘behaviour change techniques’). There has been a particular focus on how to describe active intervention components in the smallest possible ways with the development of the behaviour change technique taxonomy (BCTTv1; for example, see Cane et al., 2015).

Aspects of the BCW approach explain how intervention components work through identifying the influences, or causal mechanisms, that shape behaviour. These influences, or causal mechanisms, have been described in two complementary levels: broadly as ‘capability’, ‘motivation’ and ‘opportunity’ (i.e., the COM-B framework, where B represents behaviour); and in more detailed, with clearer traceable links to traditional health psychology theory, as the fourteen more specific domains categorised within the theoretical domains framework (TDF) (Atkins et al., 2017; Cane et al., 2012; French et al., 2012).

The BCW, when incorporating the TDF and applied to behavioural data, provides the means to identify theoretically informed intervention functions which are tailored to the context and population from which the data were collected. Furthermore, when also using the behaviour change technique taxonomy (BCTT), these broad intervention functions can be operationalised in terms of highly specific, granular, intervention components. The COM-B, intervention function and policy categories of the BCW can be seen in Figure 1.

**Figure 1.**
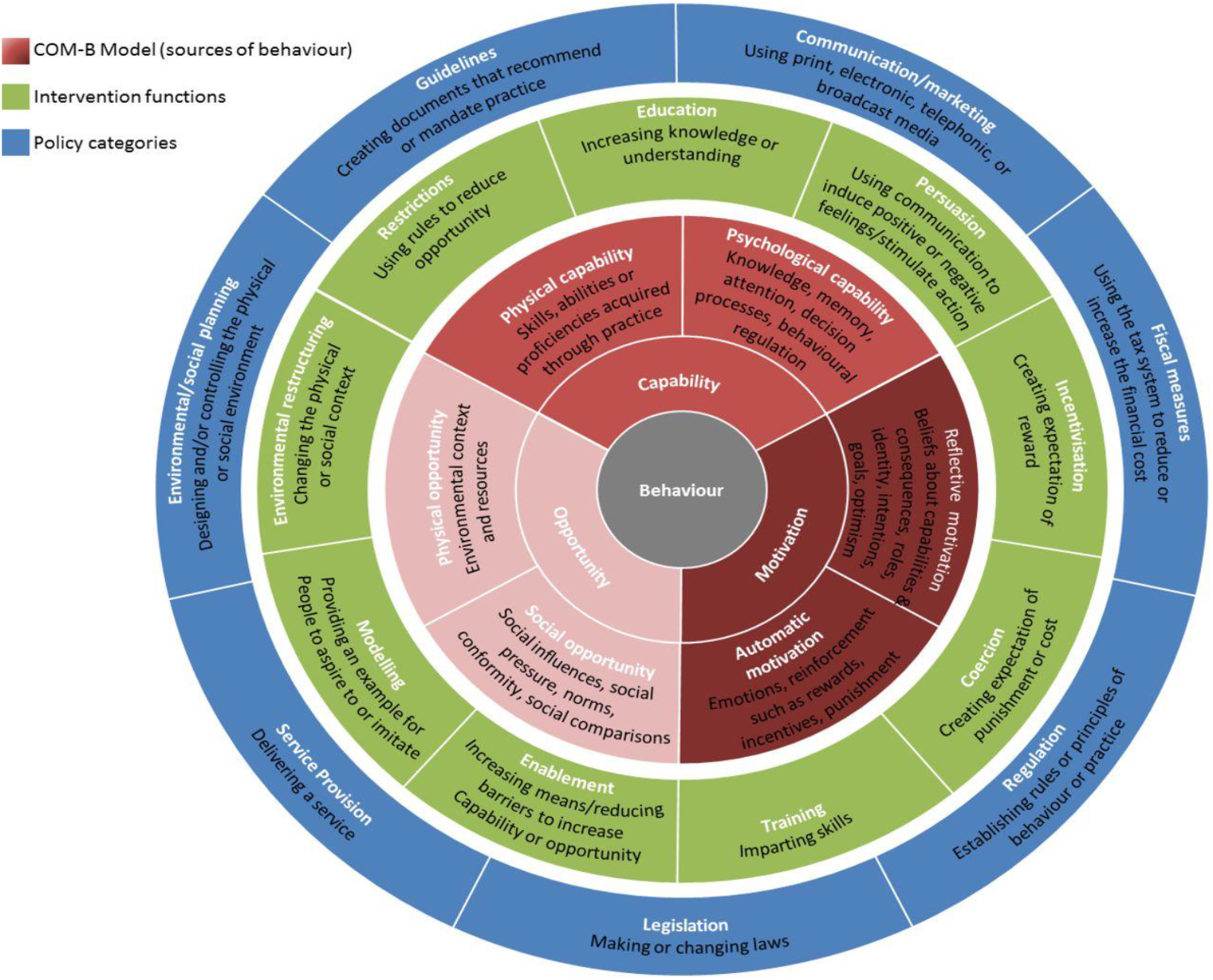
The behaviour change wheel (Michie et al., 2011). COM-B, Capability, Opportunity and Motivation Model of Behaviour *Taken from: ‘McDonagh LK, Saunders JM, Cassell J, Curtis T, Bastaki H, Hartney T, Rait G. Application of the COM-B model to barriers and facilitators to chlamydia testing in general practice for young people and primary care practitioners: a systematic review. *Implementation Science*. 2018 Dec;13(1):1-9.

### The current study

Here we report on one key element of a larger, multi-stage, multi-stakeholder intervention development process which addresses the whole of the PN process from notifying partners to accessing testing and wider care (see McQueen et al., 2020 for the larger protocol; or Flowers et al., 2021). We present our analyses of data collected, after an initial stakeholder engagement event, from heterogeneous samples of GBMSM through focus groups, followed by further stakeholder input. These focus groups were concerned with understanding the barriers and facilitators to the particular behaviour of notifying partners as the gateway step to the wider PN process. Data related to PN in general but also focused on the relational context of ‘one-off partners’ in particular. In the study, we used a variety of tools from health psychology and the behavioural sciences to assist with understanding, theorising, and suggesting ways to change these barriers in order to enhance PN for GBMSM.

### Rationale

The purpose of this study is to explore and theorise the barriers and facilitators of GBMSM notifying their ‘one-off’ sexual partners about their exposure to STIs using both the COM-B model of behaviour change and the TDF and then to make evidence informed suggestions for enhancing PN, using the BCW approach, the BCTT and stakeholder engagement.

### Research questions

1. What are the important barriers and facilitators to notifying one-off sexual partners of their exposure to STIs amongst GBMSM?
2. Which are the important COM-B factors and TDF domains when theorising important barriers and facilitators to notification?
3. Which potential intervention content can be suggested that directly addresses these important barriers and facilitators using the BCW approach?

## Methods

### Procedure

We worked closely with a range of community based organisations and informal networks to recruit a diverse sample of GBMSM from across the UK. We used social media where possible to advertise the focus groups and enable wide participation.

Potential participants were asked to read a briefing and then sign a consent from indicating their consent to partake in the study. A total of five focus groups were conducted, three in person (Glasgow, n = 4; London, n = 8; Leeds, n = 7) and two online/virtually (n = 5 and n = 4) using an interactive virtual platform (Blackboard) throughout 2020.

Focus groups lasted around two hours with two facilitators (PF and JMcQ). The focus group sessions were structured and followed a topic guide which focussed on introducing PN, a discussion of bacterial and viral STIs, an introduction to the project’s broad goals and background and then a detailed discussion of the barriers and facilitators to partner notification. White boards, sticky notes and comments in Chat functions were used to visibly record key points. The facilitators encouraged participants to talk to each other as much as possible. At the end of the focus groups, participants were provided with a debriefing.

Ethical approval was attained from Glasgow Caledonian University (HLS/NCH/19/05) and all participants provided written consent.

### Data analysis

For research question one, the transcripts from the focus groups were analysed using deductive thematic analysis to identify and prioritise barriers and facilitators to PN in GBMSM. Analysis was led by JMcQ and audited by PF.

For research question two (RQ2), the analytic tools COM-B and TDF from the behaviour change wheel (BCW) approach (Michie et al., 2014), were used to theoretically understand and explain the underlying influences/causal mechanisms of the barriers and facilitators to PN in GBMSM. Analysis was led by JMcQ and audited by PF, GV and JMcL.

For research question three, continuing use of the BCW approach, based on the COM-B and TDF elements identified and outlined in RQ2, intervention functions and BCTTs were suggested for directly addressing the barriers and facilitators to PN and thus enhancing GBMSM engagement with PN. Analysis was led by JMcQ and audited by PF, GV and JMcL.

For research question four, stakeholder engagement (representation from important third sector organisations and a range of expert healthcare professionals) was used to finalise the set of intervention suggestions. Stakeholders were asked to use the APEASE criteria (Acceptability, Practicability, Effectiveness, Affordability, Side-effects, and Equity) to accept or reject suggested intervention elements. Analysis was led by JMcQ and audited by PF.

In the section below, we provide a narrative account of the final suggested intervention components, as generated through the data and our rigorous use of the COM-B, TDF, BCW and BCTT. Our recommendations are structured into three key interrelated areas. For the interested reader, within the on-line supplementary file, we show the details of each step of our coding and present all derived recommendations.

We used these tools, in addition to stakeholder engagement (representation from important third sector organisations and a range of expert healthcare professionals), to finalise a set of intervention ideas. These ideas were all generated through the data and our rigorous use of the COM-B, TDF, BCW and BCTT.

## Results

### 1. What are the important barriers and facilitators to PN in GBMSM

In relation to the behaviour of notifying one-off sexual partners about their potential exposure to an STI, our deductive thematic analysis highlighted heterogeneous, inter-related and sometimes seemingly contradictory barriers and facilitators (See Table 1). Recruitment site, or socio-demographic factors did not explain the patterning of contradictory barriers and facilitators, suggesting a diversity of beliefs across the GBMSM population.

**Table 1.** Barriers and facilitators to notifying one-off partners amongst GBMSM

| Barriers to notifying one-off partners | Facilitators to notifying one-off partners |
| --- | --- |
| Perceptions of high levels of STI-related stigma | Perceptions of low levels of STI-related stigma |
| Perceptions of high levels of STI-related blame | Perceptions of no STI-related blame |
| Perceptions that STIs do not have serious consequences | Perceptions about the potential serious consequences of STI infections for health |
| Beliefs about the diffusion of responsibility for informing partners | Beliefs about personal responsibility for notifying partners |
| Difficulties relating to not planning to have sex with one-off partners again |  |
| Challenges in identifying anonymous partners to enable informing them |  |
| Problems with the recall of one-off partners |  |
| Difficulty finding contact details for partners met through dating apps |  |

### 2. Which are the important COM-B factors and TDF domains when theorising barriers and facilitators to notification

The barriers and facilitators to notifying partners were theorised in relation to COM-B areas and the corresponding TDF domains. Findings are presented in decreasing order of importance within the COM-B and TDF frameworks: motivation, opportunity and then capability. Within the COM-B elements, respective TDF domains are presented in relation to decreasing order of importance and those that were not important are not listed. Figure 2 visualises the overall shape of the findings with the relative size of the COM-B and TDF variables schematically representing their relative importance based on insights from the deductive thematic analysis listing important barriers and facilitators.

**Figure 2.**
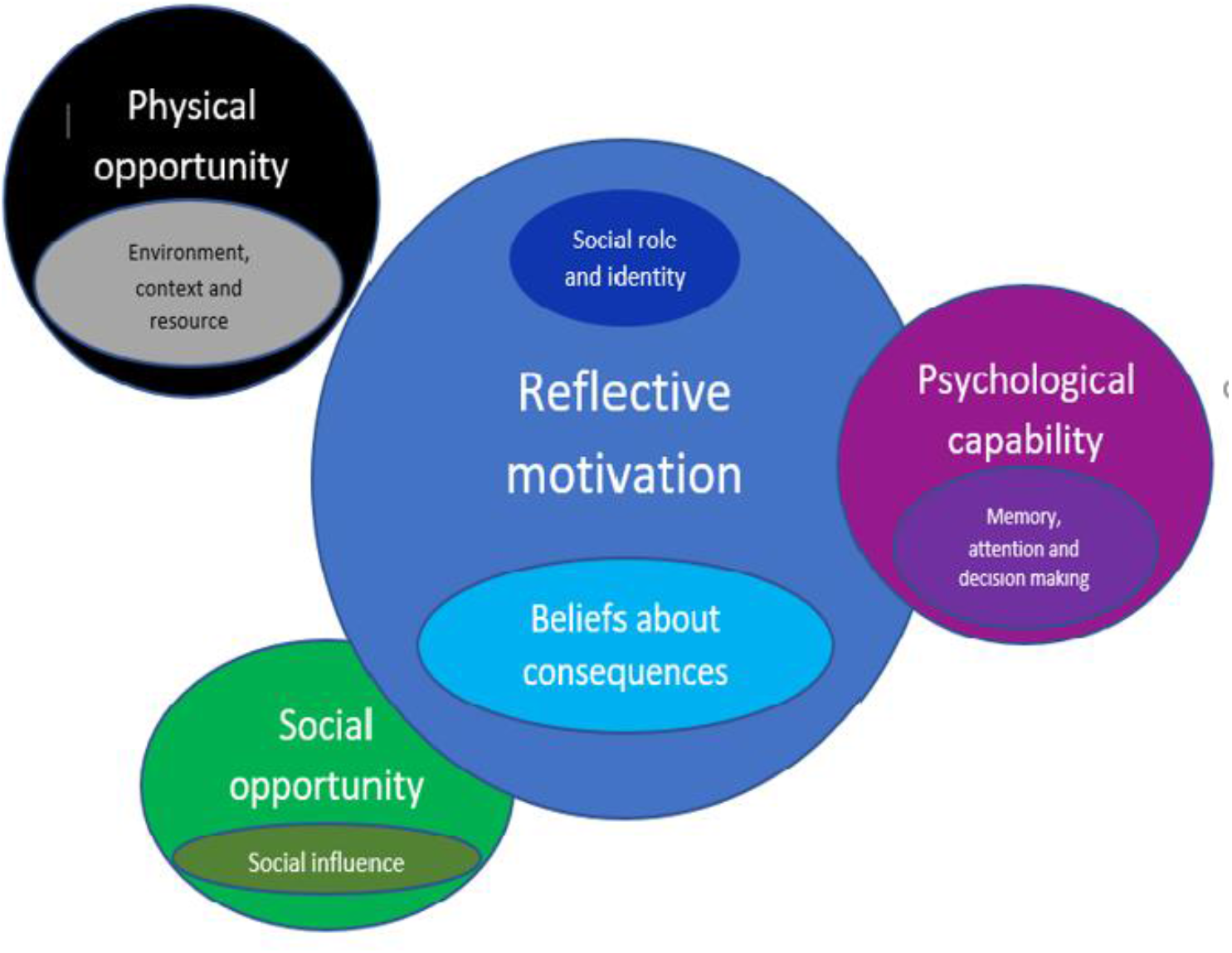
The relative importance of COM-B and TDF domains in relation to barriers and facilitators to notifying partners of their potential exposure to STIs amongst GBMSM

#### COM-B: Reflective motivation

Within the COM-B framework, the area of reflective motivation in which people make active self-conscious decisions and weigh up the costs and benefits of engaging in PN or not, was most important representing a major area of causal influence on notification behaviour. Within this, the corresponding TDF domain ‘beliefs about consequences,’ was of particular relevance. ‘*Social role and identity’* had lesser importance.

#### TDF: Beliefs about consequences

Beliefs about the consequences of notifying partners were varied and related to health, reputation, stigma and blame. These beliefs were presented as very strong influences in shaping men’s decisions to inform one-off partners of STI diagnoses.

##### Health consequences

Participants raised concerns about serious health consequences when discussing the implications of not notifying partners, reporting that: *“if it’s something like shigella, you know, hepatitis, HIV, it will have life-changing effects, or it could do anyway” (Leeds)*. Both individual and community level health implications were considered aspects that should encourage PN: “*And if that (STI) goes untreated and it gets passed around and you’ve got a massive spike, eventually the antibodies are going to stop responding, and what’s going to happen then*?” (Virtual focus group2). However, potential suicide as a result of notification was also considered a risk: “*it’s all very well to send out a message, oh, you’ve got chlamydia but if you send out a message saying you’ve got HIV, Hepatitis C, you know, that’s going to be like, whoa, someone might commit suicide*.*” (Leeds)*

##### Reputational consequences

Participants identified issues of personal reputation as shaping decisions to inform their one-off partners: *“It depends on the STI as well, but you might not want to tell them personally in case they tell other people as well or, you know, think bad of you, you end up getting a name for yourself or something like that*.*” (Leeds)*. It was considered particularly undesirable to get a reputation for chronic STIs *“…like HIV, you could end up being, oh, he’s the guy with HIV or hepatitis or something, you know*.*” (Leeds)*

##### Stigma and social consequences

The repercussions of telling partners about having possibly passed on an STI was a concern which was reinforced by admissions of expected negative feelings towards partners reporting STIs to them: “*I wouldn’t want to reconnect with someone who I think has given me chlamydia, I don’t think I’d ever want to speak to that person again. Or, they’ve given me scabies, I don’t know, it’s disgusting*.*” (virtual focus group 2); “The response you can get can vary. It can be like, oh yeah, no worries, you know, yeah, I understand, whatever. Some people have been outright, sort of, rude……as if I was like, you know…you’re carrying a plague or something*.*”* (London). On the flip-side, for individuals reporting STI discussions as routine, the element of stigma was not relevant: *“I’d say for me, (talking about STIs) it is normal, it’s not an issue. A) I’m not embarrassed, B) most of my friends are on PrEP, I’ve got friends who are HIV positive…So to me, it’s just if I get gonorrhoea, if I get chlamydia, I’m not embarrassed about it and I would tell somebody*.*” (London)*

Other beliefs about consequences related to the potential meaning of recontacting a one-off partner and the resulting social consequences: *“By contacting them, you’re having more than a one-off sexual encounter with them. You’re now having a…I thought I better tell you that I’ve got STI, so you’re starting the relationship. They’re going to go, oh yeah, let me go and get tested. Oh yeah, I’ve got an STI as well. And now you’re talking. It’s not all about the sex you’ve had, it’s finished*.*” (London)*

##### Blame

The fear of blame was an important barrier to PN and men discussed the importance of alleviating blame: *“You know, some people when you tell them that, they will automatically… they assume that you are accusing them of giving it to them”* (London). The need to consider a shared responsibility for unprotected sex was one solution offered: *“I don’t understand why they should get so emotional about it when they know the full risks … So therefore why put the blame on to just myself when it’s two people involved?” (London)*. Another suggestion was to help people *“understand that you are not generating the STI. It’s come to you …you’re not making from your body. So it’s come to you, so it’s not your fault…” (Virtual)*

#### TDF: Social role and identity

The view of lessened responsibility was identified, particularly in relation to casual partners; however, there were also opinions about the importance of notifying partners as a moral obligation to all partne*rs: “It’s about morals and I guess some people just don’t have the moral compass so they don’t care*.*” (London)*. Negative past notification experiences involving excessive emotional reactions were discussed as a barrier but also as having led to reflections on sharing the responsibility of the risk of unprotected sex and refusing to take on blame alone.

#### COM-B: Social opportunity

When we mapped the barriers and facilitators to the COM-B model, the area of social opportunity was also important in notifying. This relates to how the social world enables or constrains behaviours of interest. The corresponding TDF domain of ‘social influence’ was particularly salient.

#### TDF: Social influence

##### Norms

Beliefs about norms and responsibility featured as important in shaping PN. GBMSM model their own behaviour on the behaviour of others: *“So some people might have that attitude of, oh screw it, they didn’t tell me, so I’m not going to reciprocate. Fuck it, I’m going to deal with it and keep it moving*.*” (London)*

Elements of the subculture, such as the language used to describe STI status, were also seen as key elements of social influence that shaped notification behaviours. As one participant noted: “*consider the language as well, just like saying, you know, are you ‘clean’, it kind of adds more stigma to it, because it’s saying if you do have anything, then you’re ‘dirty’*.*” (Glasgow)*

#### COM-B: Physical opportunity

The area of physical opportunity was also found to be important within the COM-B model, with the corresponding TDF domain ‘*environmental context and resources’* being particularly pertinent. Physical opportunity relates to key features of the physical world which enable or constrain behaviours.

#### TDF: Environmental context and resource

This shaped notification behaviour primarily because it related to the dating app culture and functional elements of the dating app environment.

#### Dating app functionality

The possibility of blocking access to contact details on dating apps and deleting or changing profiles were perceived to challenge PN: *“I’ve had people on Grindr that literally…there’s a car park two doors away from my flat, they’ve driven to me, parked in the car park, we’ve shagged, they’ve left, and I’ve been blocked by the time they’ve got back in their car again. And do you know what, five days later, they’ve created a new profile, and that is what it is, but that’s where it can get quite difficult with partner notification as well*.*” (Glasgow)*

#### COM-B: Psychological capability

When we mapped the barriers and facilitators to the COM-B model, the area of psychological capability and the corresponding TDF domain of ‘*memory, attention and decision making*’ was particularly important. These kinds of causal influence relate to psychological propensities that enable or constrain behaviours.

#### TDF: Memory, attention and decision making

Memory was an important barrier to being able to notify partners about STI diagnoses, for example, not knowing who all their partners were, especially when *“on drugs”* (Leeds) or they met *“in a dark room*.*” (Leeds)*. For some, challenges with remembering all partners was also related to high numbers of diverse partners about whom the men knew very little: *“I probably couldn’t wind back my clock six months and tell you how many people I’ve slept with, I just couldn’t, in the past three months…that could be several hundred people*.*” (London)*

#### COM-B: Physical capability

Within the COM-B area of physical capability which relates to physical skills needed to engage in PN, we found no evidence of this being important.

### 3. Which potential intervention content can be suggested using the behaviour change wheel approach?

Building on our COM-B and TDF analysis of the barriers and facilitators to notifying partners, it was clear that reflective motivation and beliefs about consequences were particularly important in shaping notification behaviours. The analysis also highlighted that elements of the physical environment were important (i.e., the functionality of dating apps), aspects of social opportunity were sometimes important (i.e., social norms within the community) and one aspect of psychological capability (i.e., difficulties recalling particular partners).

The BCW approach directly builds upon COM-B and the TDF analyses to prescriptively specify tailored intervention functions for the future, generating detailed potential intervention components that can moderate modifiable barriers and facilitators to notifying partners. For example, given the relative importance of the TDFs ‘beliefs about consequences’ and to a lesser extent ‘social role and identity’, the use of the BCW highlights the potential usefulness of intervention functions ‘education’, ‘persuasion’ and ‘modelling’. Along these lines the BCW provided a clear sense of direction for tailored ways of *changing* how MSM make conscious reflective decisions about notification. The analysis also suggested, based on the other COM-B and TDF findings, that intervention functions ‘enablement’, ‘environmental restructuring’ and ‘training’ were also appropriate ways of intervening.

From the intervention functions, the BCTTv1 was then used to identify highly specific and relevant potential intervention content. In addition, stakeholders such as expert health care professionals (HCPs) and staff from non-governmental organisations (NGOs) were also consulted to ensure realistic application.

Overall, our behavioural wheel analysis suggests that to affect reflective motivation and change social opportunity, concerted effort should be made to change the norms and beliefs of GBMSM about notification. Given that we identified considerable heterogeneity in relation to the perceived consequences of notifying, a sensible way forward would be to amplify those pre-existing norms and values which already support notification. This could be accomplished through focussed efforts using the mass and social media and working to harness social influence through peer-led work. In this way, norms of notification are changed and the perceived social cost of notifying one-off partners are reduced. In turn, over time, as more notification takes place, it is possible that a positive feedback loop might reinforce notification as an anticipated injunctive and visibly descriptive social norm. Figure 3 illustrates a high-level overview of our final recommendations.

**Figure 3:**
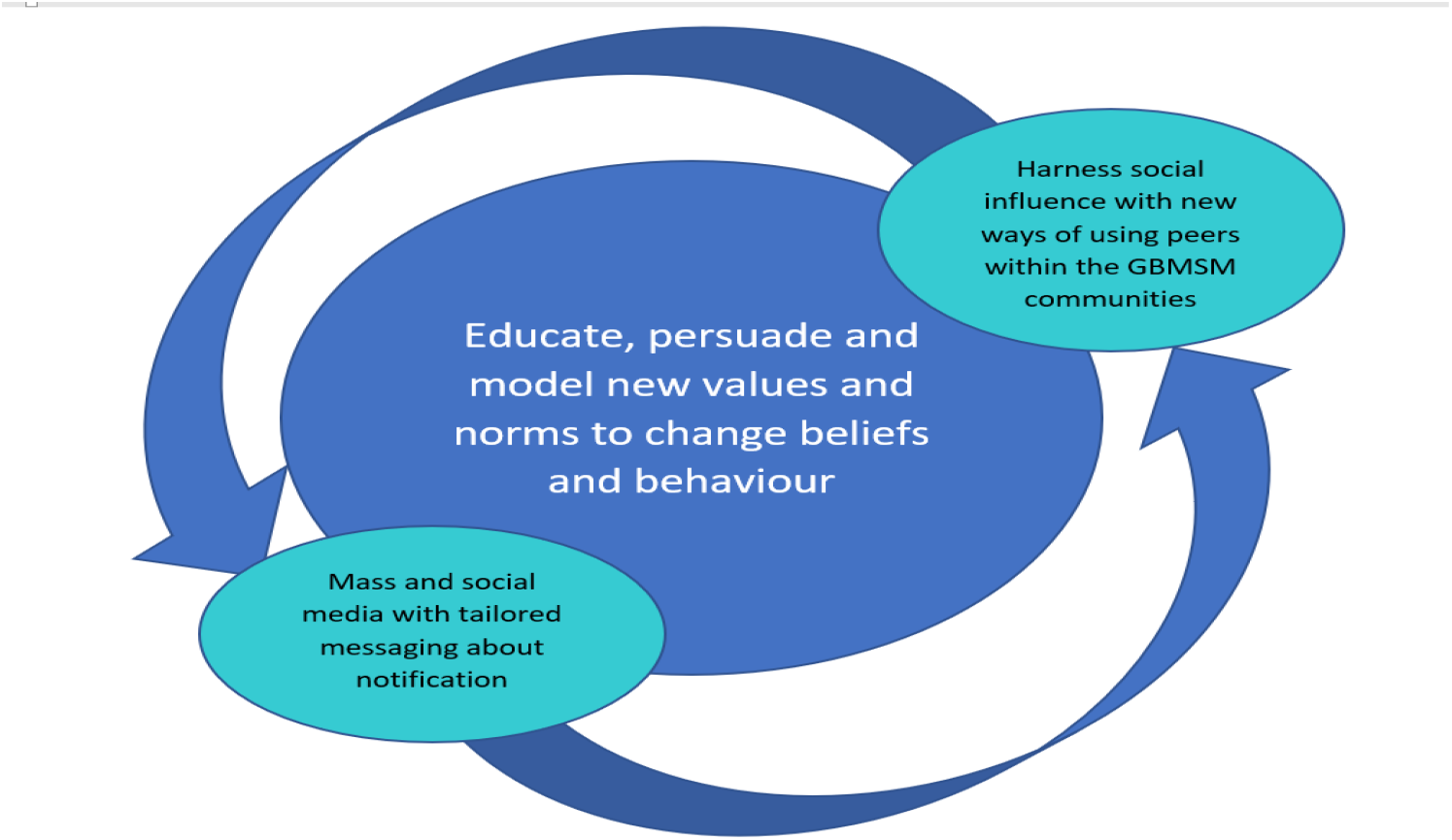
Final suggestions for approaches to intervention development to improve notification of partners of their potential exposure to STIs.

From the intervention functions, the BCTTv1 was then used to identify highly specific and relevant potential intervention content. In addition, stakeholders such as expert health care professionals (HCPs) and staff from non-governmental organisations (NGOs) were also consulted to ensure realistic application.

Overall, our behavioural wheel analysis suggests that to affect reflective motivation and change social opportunity, concerted effort should be made to change the norms and beliefs of GBMSM about notification. Given that we identified considerable heterogeneity in relation to the perceived consequences of notifying, a sensible way forward would be to amplify those pre-existing norms and values which already support notification. This could be accomplished through focussed efforts using the mass and social media and working to harness social influence through peer-led work. In this way, norms of notification are changed and the perceived social cost of notifying one-off partners are reduced. In turn, over time, as more notification takes place, it is possible that a positive feedback loop might reinforce notification as an anticipated injunctive and visibly descriptive social norm.

Our use of the BCTT within the BCW approach enabled us to go further than a broad level of suggesting future intervention content and instead suggest very granular, tailored ways in which notification of one-off sexual partners could be improved. Suggestions to alter the functionality of dating apps to enable the possibility of contact for the purpose of PN were the only main area of objection from stakeholders. For the interested reader, full details of this analysis are available within the supplementary files. In the section below, for the sake of brevity and to communicate our findings effectively, we present a higher level, narrative account of the highly specific ways our detailed analysis has suggested that notification amongst GBMSM could be improved.

### 1. Changing norms and values to change beliefs and behaviours

Intervention content should educate, persuade and model notification as normative and as part of an endorsed and preferred future culture and community value that benefits the health and wellbeing of all.

Several activities can be encouraged among GBMSM as part of this vision, namely the co-production of mass and social media intervention content and peer-led interactive approaches. Moreover, individual-level activities such as informally self-monitoring personal behaviour in relation to not stigmatising or blaming others; support and training to keep records of one-off sexual partners as a means of conducting PN if required; and participating in regular STI testing so as to facilitate the recall of more one-off partners by shortening the recall period.

Mass and social media, as well as peer-led interactive approaches, can provide potential ways of operationalising the broad principles concerned with changing the culture and norms around PN. These approaches can also provide prompts and cues in a variety of formats and locations, through key partners (e.g., HCPs, dating app providers (DAPs) and (NGOs)) to encourage GBMSM to buy into PN related activities.

In line with reducing stigma and blame and sharing responsibility, co-produced messages could assist in reframing the notification of one-off partners as being simply the ‘*right thing to do’* instead of an onerous or costly task in relation to the perceived consequences outlined earlier. Equally, messaging such as ‘what goes around comes around’ can help ensure the apparent ‘costs’ of these activities are off set with the benefits, while also making notification generally more personal than altruistic. Further, drawing on COVID experience, the possibility of a mechanism by which people using a venue (sex on premises) can provide contact details to enable PN to take place if there is an outbreak should be considered.

### 2. Mass and social media with tailored messaging about notification

Mass and social media can be powerful tools to encourage engagement notification by directly changing beliefs that STI prevention is solely an individual responsibility. Culturally sensitive messaging could be delivered through peer narratives modelling the discouraging of blaming and on-going stigmatising. Key culturally appropriate messages delivered through these media could include the presentation of key scenarios: the consequences, and contrasting outcomes, of when blame *is* and *is not* used; the possible positive and negative outcomes of STI disclosure; the possible positive outcomes associated with non-stigmatising responses to STI disclosure for individuals and communities, models and scripts of appropriate ways of notifying and responding to being notified. More complex issues could be communicated through the depiction of personal testimonies, for example, narratives showing that notification is not necessarily associated with signalling further affective or sexual commitment and instead depicting that it can feel good to engage in notification by contributing to everyone’s health.

### 3. Peer-led work and its potential content

As part of a range of intervention formats, peer education can provide invaluable support to normalising responsibility for others and highlighting the expectations and consequences of being responsible for others within communities. Intervention materials, or content, ideally co-produced by peers, NGO staff and HCPs, and delivered by peers, could be persuasive, starting as a source of support at point of diagnosis and continuing to foster a culture of peer support to further help increase confidence and empower GBMSM to talk about STIs and engage in PN so it can become normalised.

Messaging, or interactional content, could focus on the health consequences and wider impacts of undiagnosed STIs, model possible negative consequences and include using techniques like anticipated regret to encourage men to consider how they might feel about someone close to them becoming ill as a result of undiagnosed syphilis. Peer-led interventions may be particularly useful in addressing a range of interactional and thus social barriers to notification. Peer-led intervention elements are a visible depiction of community norms. In this way, peer-education can provide invaluable support in normalising the responsibility of individuals diagnosed with an STI to inform others and highlighting the expectations and consequences of being responsible for others within communities. Intervention materials/content would ideally be co-produced by peers, NGO staff and HCPs, and delivered by peers. This could be persuasive, act as a source of support, and continue to foster a culture of peer support to further help increase confidence and empower GBMSM to talk about STIs and engage in PN so it can become normalised.

## Discussion

Using theory and a series of tools designed to generate specific, tailored, intervention components we have suggested a clear direction of travel for future intervention. Our suggestions make the most of the reported facilitators to notification yet also attempt to directly reduce reported barriers. Our long list of potential recommendations were appraised and reduced by various stakeholders. While our findings support previous research on barriers to notification such as fear and shame relating to STIs, and concern about negative reactions from partners (Balaji et al., 2017), here we have been able to move the field further in relation to suggestions for future intervention development.

With regards to our first research question concerning important barriers and facilitators to notifying partners, we found these related to the perceived social consequences of notification in relation to social aspects of the lives of GBMSM (e.g., STI-related stigma, perceptions of blame) but also in relation to perceived health consequences (e.g., the severity of STIs). Critically, there was noticeable heterogeneity of these beliefs across the sample and we took this to reflect a heterogeneity of beliefs across the wider MSM communities. As well as the pressing weight of these psychosocial barriers, we also noted a series of practical barriers to notifying one-off partners including problems with recall of one-off partners and further practical challenges of contactability such as the functionality of dating apps which may have facilitated men meeting for one-off sex in the first place. The key stakeholder objections to our recommendations lay in making functionality changes to the DAPs to ensure some contact for PN is possible.

In relation to our second research question, here using the conceptual language of COM-B, we theorised these barriers and facilitators and found that they related primarily to reflective motivation (i.e. the weighing up of costs and benefits of notification), yet also and relatedly to social opportunity (the way the social environment constrains or enables notification to occur). Thus, it appears important causal influences relate to the psychosocial and cultural context of MSM and their varied communities. Key expectations, assumptions, norms, values, identities and other social influences were important in shaping the way men weighed up the costs and benefits of notifying one-off partners of their potential exposure to STIs. Critically, in relation to moving the field forwards, these are relatively easily modifiable through community, peer and individual level intervention. Of lesser importance than these issues we noted that other barriers and facilitators could be theorised in relation to physical opportunity (i.e. external world) and psychological capability.

In relation to our third research question, we found key intervention functions useful to improve notification to be primarily ‘education’, ‘persuasion’ and ‘modelling’. To a lesser extent the use of ‘enablement’, ‘environmental restructuring’ and ‘training’ were also relevant. Using mass and social media, along with peer-led work, to encourage change in the culture of PN among GBMSM, norms and values may be shifted towards a focus on the wider benefits of discouraging blame and stigma. If STI blame and stigma are discouraged and notification is encouraged as the beneficial choice, both from a personal and community perspective, detailed messaging, narratives and modelling may facilitate the normalisation of STI discussions and notification.

### What have we uniquely done here?

In the current paper we have used behavioural theory and associated theoretical frameworks to supply a systematic process to strengthen intervention development (Gould et al., 2014; Michie et al., 2014). In this way, our analyses move the field forwards from simple descriptions of barriers and facilitators to notification, and instead provide detailed and concrete ways of intervening. These approaches have been used to improve various health outcomes such as increasing chlamydia testing in primary care (McDonagh et al., 2020) and enhancing nurses’ use of electronic medication management systems (Debono et al., 2017).

## Limitations

Study limitations include our reliance on qualitative data only to understand the barriers and facilitators to notification amongst GBMSM and their one-off partners. If resource had permitted, we would have conducted more comprehensive mixed methods research to achieve a firmer grasp of the population-level patterning of barriers and facilitators. Equally our reliance on convenience sampling for recruiting diverse GBMSM also represents a potential source of bias and while our original intention had been to recruit GBMSM from a range of geographical areas the impact of COVID restrictions reduced our ability to do so.

## Conclusions

The combination of the TDF and the BCW approach, including the COM-B model, provided a useful theoretical framework for examining the barriers and facilitators to PN regarding STI diagnosis for one-off partners among GBMSM. Further, the BCW approach also provided a framework for identifying intervention functions and BCTs to develop specific and targeted intervention components to overcome the identified barriers. We presented theory driven suggestions for tailored interventions to address specific barriers to PN, including (1) mass and social media messages to reduce blame and stigma around STIs and encourage individual and shared responsibility to engage in PN through education, persuasion and modelling; (2) health information pages on dating apps to encourage regular testing and highlight the benefits, ease and speed of PN; and (3) peer-led education, co-created with NGOs and HCPs, to further reduce stigma, increase personal responsibility and normalise PN.

We have also provided useful practical insights and methodological contributions for future research around intervention development. The growing literature on intervention development that is underpinned by theoretical models will ideally help address ongoing health challenges such as notification, while also establishing strong evidence for associated new interventions such as contact tracing for COVID patients. This should lead to the detailed examination of the more and less successful elements of interventions, the further development of new interventions followed by implementation and testing.

## Supporting information

GUIDED checklist

## Data Availability

Data are available from the study authors upon reasonable request

## Online Supplementary File

| Barriers to enacting this intervention component | Facilitators to enacting this intervention component | TDF Domain | COM-B | Intervention function (BCW) | Potential behaviour change techniques | What does the analysis tell us about potentially useful ways forwards in relation to potential intervention content?<br>Colour coding reflects the end results of our work with stakeholders and the APEASE criteria (green keep, amber -reconsider and red-abandon) |
| --- | --- | --- | --- | --- | --- | --- |
| GBMSM find it difficult to notify their partners because of STI-related stigma | GBMSM find it easier to notify their partners because there is little STI-related stigma | Social influences | Social Opportunity | Education<br>Persuasion<br>Enablement<br>Modelling | 11.2 *Reduce negative emotions*<br>5.3 *Information about social and environmental consequences*<br>5.6 *Information about emotional consequences*<br>5.1 *Information about health consequences*<br>2.3 *Self-monitoring of behaviour*<br>4.1 *Instructions on how to perform a behaviour*<br>6.1 *Demonstration of a behaviour*<br>9.3 *Comparative imaginings of future outcomes*<br>16.3 *Vicarious consequences*<br>3.2 *Social support practical*<br>12.5 *Adding objects to environment* | Our initial exploration of the broad behavioural domain of disclosing an STI diagnosis suggests that in order to moderate beliefs about negative consequences (11.2), intervention approaches that <u>educate</u> (12.5) and <u>persuade</u> all men about STI stigma (5.3, 5.1, 5.6) and provide feedback on its consequences (5.1, 5.3, 5.6) to other men may be a good way to systematically reduce these perceived barriers and enable more men to disclose without fear of negative sequelae. (16.3, 11.2, 5.3, 5.6, 9.3)<br>To <u>enable</u> men to inform their partners it may be possible to <u>model</u> examples of disclosing an STI within future intervention content (6.1, 4.1), and encourage men to informally self-monitor their own behaviour in relation to stigmatising others (for example, self-checking their correspondence on dating apps). (2.3, 5.6)<br>Intervention approaches, whether delivered within mass or social media, or through face to face interaction, could contrast examples of where disclosure of STI diagnosis has resulted in good or bad outcomes. (16.3, 9.3, 5.6, 5.3)<br>Interventions could be delivered <u>in a range of formats</u> including mass and social media (12.5), potential peer education or the provision of support materials at point of diagnosis (e.g., how to overcome barriers associated with perceived stigma and engage in PN, particularly with one off partners) (3.2)<br>Intervention ideas like this could <u>foster a culture of peer support</u> (3.2, 11.2) to help increase confidence and empower GBMSM to talk about STIs and engage in PN. In this way PN can become normalised. (3.2) |

| Barriers to enacting this intervention component | Facilitators to enacting this intervention component | TDF Domain | COM-B | Intervention function (BCW) | Potential behaviour change techniques | What does the analysis tell us about potentially useful ways forwards in relation to potential intervention content? |
| --- | --- | --- | --- | --- | --- | --- |
| GBMSM find it difficult to notify their partners because of perceived STI-related blame | GBMSM find it easier to notify their partners because there is no perceived STI-related blame | Beliefs about consequences | Reflective motivation | Education<br>Persuasion<br>Modelling | 5.3 Information about social and environmental consequences<br>11.2 Reduce negative emotions<br>5.2 Salience of consequences<br>5.6 Information about emotional consequences<br>5.1 Information about health consequences<br>12.2 Restructuring the social environment<br>9.1 Credible Source<br>6.1 Demonstration of a behaviour<br>6.3 Information about others approval<br>5.5 Anticipated regret<br>9.3 Comparative imaginings of future outcomes<br>16.3 Vicarious consequences | Future intervention content aimed at men should directly <u>address the issue of perceived blame</u> (11.2) as it is central barrier to engaging with PN. (5.3, 5.1, 5.2)<br>Intervention messaging within social and/or mass media could focus on key organisations (e.g., NGOs, DAPs) (9.1) <u>endorsing</u> the problematisation of STI-related blame. (9.1, 5.2, 5.1, 5.3, 5.6)<br>Culturally sensitive messaging could <u>model</u> non-stigmatising responses to STI disclosure (11.2, 6.1) and <u>enable</u> the viewer to compare their typical reaction to that which is depicted. Together these would help generate a non-stigmatising culture amongst men. (5.3, 5.1, 6.3)<br>Messaging within media materials could also use narratives to <u>persuade</u> men to change their blaming and stigmatising (11.2, 5.1, 5.6) by <u>showing the consequences of their blaming and not blaming</u> by showing that blame can lead to a lack of disclosure (5.2) which in turn could result in the stigmatiser becoming infected with an STI because the role of perceived stigma and blame prevented men from timely disclosure of an STI. (5.2, 5.3, 6.3)<br>A sliding doors scenario may be helpful (9.3, 16.3, 5.1, 5.6) to depict the comparison of blame vs non-blame and detail the varied consequences (emotions, health, infection rates staying high within the communities). (5.5, 12.2, 6.1) |
| GBMSM find it difficult to notify partners because they believe it's not their responsibility | GBMSM find it easier to notify partners because they believe it's their responsibility | Social role and identity | Reflective motivation | Education<br>Persuasion<br>Modelling | 5.3 Information about social and environmental consequences<br>5.1 Information about health consequences<br>7.1 Prompts and Cues<br>12.2 Restructuring the social environment<br>9.1 Credible Source<br>6.1 Demonstration of a behaviour<br>12.5 Adding objects to the environment<br>6.2 Social comparison<br>6.3 Information about others approval | Working with key partners such as NGOs, Public health or DAPs, (9.1) intervention messaging could focus directly on <u>educating</u> men about the issue of perceived responsibility and highlight that there are diverse perspectives on it amongst men, and highlight responsibility (5.1, 5.3) for others as part of an endorsed and preferred future culture and communities that will benefit the health and wellbeing of all. (5.3, 9.1)<br>This messaging could also focus on <u>modelling</u> the depiction the diverse negative consequences (5.3, 5.1) of believing that its every *individuals* responsibility to deal with their STI only and not engage in PN. (5.1, 5.3, 6.2)<br>Messaging could include interactive diagram to illustrate how one infected, sexually active individual can infect a number of others without knowing and <u>persuading</u> men of shared responsibility (12.5, 5.3)).<br>Messaging could change the social environment (12.2) and <u>model</u> the behaviour of informing one-off partners and include testimonials and personal accounts (9.1, 6.3) of how good it is to let others know, and how good it feels to contribute to everyone's health. (6.3, 5.1, 7.1,)<br>Intervention materials, or intervention content, if delivered by <u>peers</u>, or within key interactions between NGO staff and men (9.1), could be <u>persuade and model informing one-off partners</u> (6.1) clearly normalising responsibility for others, expectation and consequence of being responsible for others within communities. Ideally such messaging would be co-produced by peers, NGO staff and HCPs. (9.1, 6.1, 12.5, 5.1, 5.3) |
| Barriers to enacting this | Facilitators to enacting this | TDF Domain | COM-B area | Intervention function (BCW) | Potential behaviour change techniques | What does the analysis tell us about potentially useful ways forwards in relation to potential intervention content? |

| intervention component | intervention component |  |  |  |  |  |
| --- | --- | --- | --- | --- | --- | --- |
| GBMSM find it difficult to notify partners because they don't perceive the consequences of STIs to be serious | GBMSM find it easier to notify partners because they perceive the consequences of STIs to be serious | Beliefs about consequences | Reflective motivation | Persuasion<br>Modelling<br>Education | 5.1 Information about health consequences<br>5.2 Salience of consequences<br>5.3 Information about social and environmental consequences<br>9.1 Credible Source<br>6.1 Demonstration of a behaviour<br>6.3 Information about others approval<br>5.5 Anticipated regret<br>9.3 Comparative imaginings of future outcomes | Information messaging, or the content of interactive interventions such as peer-led work, (9.1) or interactions between NGO or HCP staff, could focus on <u>educating</u> men (5.1, 5.3, 6.3, 6.1) about the important health consequences and wider impacts of undiagnosed STIs to <u>persuade</u> men to disclose STI diagnoses. (5.2)<br>Messaging, or interactional content, could <u>model</u> consequences and include using BCTs such as anticipated regret (5.5) to <u>persuade</u> the viewer how they might feel about someone close to them becoming ill as a result of undiagnosed syphilis. Messaging along the lines of 'what goes around, comes around' may be helpful to make this issue more personal than altruistic (9.3). |
| Men find it difficult to notify one-off partners because they don't want to engage with them again |  | Beliefs about consequences | Reflective motivation | Education<br>Persuasion<br>Modelling | 5.1 Information about health consequences<br>5.3 Information about social and environmental consequences<br>5.2 Salience of consequences<br>9.1 Credible Source<br>6.3 Information about others approval<br>13.2 Framing/ reframing<br>7.1 Prompts cues | Information messaging, or the content of interactive interventions such as peer-led work, or interactions between NGO or HCP staff, could <u>educate</u> and <u>persuade</u> men to engage to inform one off partners by <u>modelling</u> the varied consequences (5.1, 5.2, 5.3) of non-disclosure and the idea that what goes around comes around. (e.g. consider visuals to depict consequences in relation to onwards infection or consider communicating cost to the NHS). (5.2)<br>Co-produced messaging (with peers, NGOs and HCPs) about the need to do PN (9.1) and overcome intentions not to do PN with one off partners should be branded to show clear endorsement from trusted organisations (e.g. NGOs) that signal approval for this kind of PN. (6.3)<br>Co-produced messaging (with peers, NGOs and HCPs) (9.1) about reframing informing one off partners as not being a big deal but as being the simple 'right thing to do' (13.2, 6.3).<br>Key partners such as STI clinics and NGOs (9.1) use motivational interviewing skills (5.1, 5.3, 5.2) to encourage GBMSM to inform more one-off partners providing prompts and cues to promote optimal partner notification behaviour (7.1). |

| Barriers to enacting this intervention component | Facilitators to enacting this intervention component | TDF Domain | COM-B area | Intervention function (BCW) | Potential behaviour change techniques | What does the analysis tell us about potentially useful ways forwards in relation to potential intervention content? |
| --- | --- | --- | --- | --- | --- | --- |
| GBMSM find it difficult to notify their one-off partners because they do not know who they have had sex with |  | Memory attention and decision process | Psychological capability | Environmental restructuring | 12.2 Restructuring the social environment<br>12.1 Restructuring the physical environment<br>12.5 Adding objects to the environment | Drawing on COVID experience, consider a mechanism by which people using a venue (sex on premises) provide contact details to enable PN to take place if there is an outbreak (wider stakeholders) (12.1, 12.2, 12.5) |
| GBMSM find it difficult to notify their one-off partners due to issues with recall associated with high numbers of sexual partners |  | Memory attention decision process | Psychological capability | Training<br>Environmental restructuring<br>Enablement | 2.3 Self-monitoring of behaviour<br>3.2 Social support practical<br>9.1 Credible source<br>5.2 Salience of consequences<br>12.5 Adding objects to the environment<br>8.3 Habit formation<br>4.4 Behavioural experiment<br>7.1 Prompts/Cues<br>1.1 Goal setting (behaviour) | Work with key partners to support and train men to keep records of their one-off sexual partners as a means of conducting PN if required. (2.3)<br>Restructure the social environment by connecting record keeping to wider understandings of PN and PN related education devised by NGOs, public health (9.1) 'what goes around comes around' (5.2) to ensure the apparent 'costs' of this activity are off-set with the apparent benefits (e.g. you will be told if one of your one off partners has an STI because they have kept a record) (8.3, 1.1)<br>Beyond supporting men, and chiming with current policy, consider ways to encourage GBMSM to participate in regular STI testing (8.3, 7.1) as they are then <u>enabled</u> to recall more one-off partners due to a shorter partner recall period (3.2).<br>Consider the use of prompts and cues (7.1) in a variety of formats and locations i.e. digital, online, written prompts in clinics, on web-pages (12.5) and NGO premises from key partners such as HCPs, DAPs, NGOs (9.1) to enable GBMSM to do PN (3.2, 8.3, 4.4) |
| **GBMSM find it difficult to notify one-off partners met on dating apps as the app can block access to contact details meaning it is not possible for GBMSM to reconnect with one-off partners** |  | Environmental context and resources | Physical opportunity | Environmental restructuring<br>Enablement | 12.1 *Restructuring the physical environment*<br>12.2 *Restructuring the social environment*<br>12.5 *Adding objects to the environment*<br>5.1 Information about health consequences<br>5.3 Information about social and environmental consequences<br>3.2 Social support practical<br>6.3 Information about others approval | Working directly with DAPs <u>restructure</u> dating apps (12.1) to offer a function to allow GBMSM to contact their one-off partners who have blocked them (12.2)<br>Working directly with DAPs <u>restructure</u> dating apps to offer a function to allow GBMSM to contact their one-off partners with deleted profiles for PN purposes (12.5)<br>Dating apps offer a dedicated sexual health info/ awareness page which users are prompted to access before accessing their profile (12.5, 12.1) This could a) raise awareness for the need of regular testing, especially after one-off, anonymous and unprotected sex, and b) highlight the benefits, ease and quickness of informing any partner including one-offs of a possible STI . (5.1, 5.3).<br>Highlighting that there are pre-scripted text messages (12.5, 3.2). Providing figures on x number of one-off sex partners having been notified, personal and NHS costs saved by notifying one-offs, (5.3) and personal testimonies that there are no further obligations such as forming a relationship (if the person does not want to), i.e. people just want to be told that they might have an STI (6.3). |

## Funding acknowledgement

This work presents independent research funded by the National Institute for Health Research (NIHR) under its Programme Grants for Applied Research Programme (reference number RP-PG-0614-20009).

The views expressed are those of the author(s) and not necessarily those of the NIHR or the Department of Health and Social Care. The funders had no role in study design, collection, management, analysis and interpretation of data; writing of the report and the decision to submit the report for publication.

