## Supplementary material for "How can we increase notification of exposure to STIs between gay and bisexual men?: intervention development using stakeholder engagement, qualitative methods and behavioural science": GUIDED checklist

**GUIDED – a guideline for reporting for intervention development studies.**

Supplementary File 1: Blank Checklist

| Item description | Explanation | Page in manuscript where item is located | Other* |
|---|---|---|---|
| 1. Report the context for which the intervention was developed. | Understanding the context in which an intervention was developed informs readers about the suitability and transferability of the intervention to the context in which they are considering evaluating, adapting or using the intervention. Context here can include place, organisational and wider socio-political factors that may influence the development and/or delivery of the intervention (15). | p4-5 | |
| 2. Report the purpose of the intervention development process. | Clearly describing the purpose of the intervention specifies what it sets out to achieve. The purpose may be informed by research priorities, for example those identified in systematic reviews, evidence gaps set out in practice guidance such as The National Institute for Health and Care Excellence or specific prioritisation exercises such as those undertaken with patients and practitioners through the James Lind Alliance. | p 5-6 | |
| 3. Report the target population for the intervention development process. | The target population is the population that will potentially benefit from the intervention – this may include patients, clinicians, and/or members of the public. If the target population is clearly described then readers will be able to understand the relevance of the intervention to their own research or practice. Health inequalities, gender and ethnicity are features of the target population that may be relevant to intervention development processes. | p6 | |
| 4. Report how any published intervention development approach contributed to the development process | Many formal intervention development approaches exist and are used to guide the intervention development process (e.g. 6Squid (16) or The Person Based Approach to Intervention Development (17)). Where a formal intervention development approach is used, it is helpful to describe the process that was followed, including any deviations. More general approaches to intervention development also exist and have been categorised as follows (3):- Target Population-centred intervention development; evidence and theory-based intervention development; partnership intervention development; implementation-based intervention development; efficacy-based intervention development; step or phased-based intervention development; and intervention-specific intervention development (3). These approaches do not always have specific guidance that describe their use. Nevertheless, it is helpful to give a rich description of how any published approach was operationalised | p6-9 | |
| 5. Report how evidence from different sources informed the intervention development process. | Intervention development is often based on published evidence and/or primary data that has been collected to inform the intervention development process. It is useful to describe and reference all forms of evidence and data that have informed the development of the intervention because evidence bases can change rapidly, and to explain the manner in which the evidence and/or data was used. Understanding what evidence was and was not available at the time of intervention development can help readers to assess transferability to their current situation. | p 6-9 | |
| 6. Report how/if published theory informed the intervention development process. | Reporting whether and how theory informed the intervention development process aids the reader's understanding of the theoretical rationale that underpins the intervention. Though not mentioned in the e-Delphi or consensus meeting, it became increasingly apparent through the development of our guidance that this theory item could relate to either existing published theory or programme theory | p 6-9 | |
| 7. Report any use of components from an existing intervention in the current intervention development process. | Some interventions are developed with components that have been adopted from existing interventions. Clearly identifying components that have been adopted or adapted and acknowledging their original source helps the reader to understand and distinguish between the novel and adopted components of the new intervention. | N/A | |
| 8. Report any guiding principles, people or factors that were prioritised when making decisions during the intervention development process. | Reporting any guiding principles that governed the development of the application helps the reader to understand the authors' reasoning behind the decisions that were made. These could include the examples of particular populations who views are being considered when designing the intervention, the modality that is viewed as being most appropriate, design features considered important for the target population, or the potential for the intervention to be scaled up. | p 6-9 | |

1

| Item description | Explanation | Page in manuscript where item is located | Other* |
|---|---|---|---|
| 9. Report how stakeholders contributed to the intervention development process. | Potential stakeholders can include patient and community representatives, local and national policy makers, health care providers and those paying for or commissioning health care. Each of these groups may influence the intervention development process in different ways. Specifying how differing groups of stakeholders contributed to the intervention development process helps the reader to understand how stakeholders were involved and the degree of influence they had on the overall process. Further detail on how to integrate stakeholder contributions within intervention reporting are available (19). | p 6, 8, 9, 15, 16 | |
| 10. Report how the intervention changed in content and format from the start of the intervention development process. | Intervention development is frequently an iterative process. The conclusion of the initial phase of intervention development does not necessarily mean that all uncertainties have been addressed. It is helpful to list remaining uncertainties such as the intervention intensity, mode of delivery, materials, procedures, or type of location that the intervention is most suitable for. This can guide other researchers to potential future areas of research and practitioners about uncertainties relevant to their healthcare context. | p 19, 20 | |
| 11. Report any changes to interventions required or likely to be required for subgroups. | Specifying any changes that the intervention development team perceive are required for the intervention to be delivered or tailored to specific sub groups enables readers to understand the applicability of the intervention to their target population or context. These changes could include changes to personnel delivering the intervention, to the content of the intervention, or to the mode of delivery of the intervention. | p 19, 20 | |
| 12. Report important uncertainties at the end of the intervention development process. | Intervention development is frequently an iterative process. The conclusion of the initial phase of intervention development does not necessarily mean that all uncertainties have been addressed. It is helpful to list remaining uncertainties such as the intervention intensity, mode of delivery, materials, procedures, or type of location that the intervention is most suitable for. This can guide other researchers to potential future areas of research and practitioners about uncertainties relevant to their healthcare context. | p 16-20 | |
| 13. Follow TIDieR guidance when describing the developed intervention. | Interventions have been poorly reported for a number of years. In response to this, internationally recognized guidance has been published to support the high quality reporting of health care? interventions[5]and public health interventions[14]. This guidance should therefore be followed when describing a developed intervention. | N/A | |
| 14. Report the intervention development process in an open access format. | Unless reports of intervention development are available people considering using an intervention cannot understand the process that was undertaken and make a judgement about its appropriateness to their context. It also limits cumulative learning about intervention development methodology and observed consequences at later evaluation, translation and implementation stages. Reporting intervention development in an open access (Gold or Green) publishing format increases the accessibility and visibility of intervention development research and makes it more likely to be read and used. Potential platforms for open access publication of intervention development include open access journal publications, freely accessible funder reports or a study web-page that details the intervention development process. | we will be aiming for an open access publication | |

*e.g. if item is reported elsewhere, then the location of this information can be stated here.



Duncan E, *et al. BMJ Open* 2020; 10:e033516. doi: 10.1136/bmjopen-2019-033516
